# The Impact of Laterality on the Incidence and Prognosis of Epithelial Ovarian Cancer

**DOI:** 10.1101/2023.08.26.23294664

**Authors:** Yang Zhang, Yucong Huang, Jihui Kang, Shuzhong Yao, Langyu Gu, Guofen Yang

## Abstract

Epithelial ovarian cancer (EOC) is the most common type of ovarian cancer, and its mortality rate is the highest among gynecological malignancies. Despite numerous factors being linked to the prognosis of EOC, the impact of ovarian laterality has received limited attention. In this study, we comprehensively examined the effects of laterality (left-right and bilateral-unilateral) on the incidence and prognosis of EOC, with a particular focus on different subtypes. By utilizing a large clinical database, we found that laterality differences primarily existed between unilateral and bilateral cases in terms of both incidence and prognosis. Specifically, unilateral tumour development was predominantly observed in patients with clear cell, endometrioid, and mucinous ovarian cancer subtypes, while bilateral involvement was more common in serous ovarian cancer. Laterality differences, reflecting disparities between the left and right sides, were primarily observed in various stages of the overall population and within specific EOC subtypes. Specifically, significant differences in EOC incidence between the left and right sides at different stages were observed in the overall population, as well as in clear cell, endometrioid, and serous ovarian cancer subtypes. Although no significant differences in the incidence rate between the left and right sides were noted for mucinous ovarian cancer, the prognosis was substantially better on the right side compared to the left side. These findings underscore the importance of considering ovarian laterality, both in terms of left-right and bilateral-unilateral aspects, as a critical factor associated with the incidence and prognosis of EOC. Therefore, it should be taken into account in clinical practice, particularly in the context of different tumour stages and subtypes of EOC.

## Introduction

Ovarian cancer is a significant concern in the field of gynaecology, with epithelial ovarian cancer (EOC) being the most common type^1,2^. It accounts for around 85% to 90% of all ovarian malignant tumours^3^. Despite advancements in treatment, the 5-year survival rate for EOC remains below 50%, posing a serious threat to women’s health^4^. The standard treatment plan involves surgery and chemotherapy, followed by targeted therapy^5^. However, the prognosis of EOC patients can vary greatly due to the strong individual heterogeneity among patients^6^. Therefore, it is crucial to improve the diagnosis and prognosis of EOC in order to develop precision therapies for patients.

The prognosis of EOC is influenced by various factors including age, tumour grade and stage, histological type, and treatment^7–9^. However, the impact of laterality, or the side of the ovary affected by cancer, on EOC prognosis is often overlooked. Some symmetrical organs affected by cancer, such as breast, testicles, and lungs, have shown different prognosis outcomes depending on the side affected. For example, patients with left testicular cancer, right lung cancer, and right breast cancer have been found to have better survival rates compared to those with contralateral disease^8,10–12^. Even among benign ovarian tumours, the incidence of mature teratoma varies significantly between the left and right sides^13^. However, few studies have analyzed the impact of ovarian laterality on incidence and prognosis of EOC. Existing limited studies have only compared the prognosis between unilateral and bilateral factors, without considering the differences between the left and right sides^14,15^. Additionally, EOC can be divided into different subtypes, including endometrioid, clear cell, serous and mucinous ovarian cancers, with unique molecular pathways, clinic treatment, and survival rates^16^. The prognoses of these histological subtypes are thus cannot be generalized^17–21^. Therefore, it remains unclear whether laterality should be considered as an effective factor for incidence and prognosis of EOC, especially for different subtypes^22–24^.

In this study, our aim is to utilize existing clinical databases to investigate the impact of ovarian laterality (both unilateral-bilateral and left-right) on the incidence and prognosis of EOC. Specifically, we intend to conduct a comprehensive analysis focusing on different histological subtypes of EOC. We hope to provide valuable insights into the prognostic significance of laterality in EOC and its implications for personalized treatment strategies.

## Materials and Methods

### 1. Population

The study population consisted of patients diagnosed with EOC between 2010 and 2017 from the SEER (Surveillance, Epidemiology, and End Results) Research Plus database, 18 Registries, Nov 2020 sub (2000-2018). The data was obtained using SEER*Stat software (version 8.4.1.2) (https://seer.cancer.gov/seerstat/).

The inclusion criteria for the study were as follows: i) patients with a pathological diagnosis of EOC based on the International Classification of Diseases for Oncology, Third Edition (ICD-O-3) anatomical code (C56.9) and morphological codes (8310/3, 8313/3, 8380/3, 8381/3, 8382/3, 8383/3, 8441/3, 8460/3, 8461/3, 8470/3, 8471/3, 8480/3, 8481/3, and 8482/3); ii) availability of complete information on histological type, EOC stage (according to the seventh edition of the AJCC^25^ or SEER combination stage^26^), and laterality; iii) active follow-up to accurately record the cause of death and survival time. The exclusion criteria included cases where EOC confirmation through cytological or histological means was lacking, as well as instances where grade classification and race information were incomplete.

The analysis was conducted at two different levels: i) the overall population, which included 20,760 patients with EOC; ii) the four histological subtypes: clear cell ovarian cancer (N=1567), endometrioid ovarian cancer (N=3422), serous ovarian cancer (N=14145), mucinous ovarian cancer (N=1287). Details about the sampling information can be found in Figure 1.

### 2. Variables

The demographic and clinical characteristics of patients were defined as follows: age (<60, ≥60), ethnicity (white, black, American Indian/Alaska Native AI and Asian or Pacific Islander API), laterality (left, right and bilateral), FIGO stage (I II III and IV), tumour grade (well, moderately, poorly and undifferentiated), and whether there were chemotherapy and radiotherapy (Figure 1).

**Figure 1.**
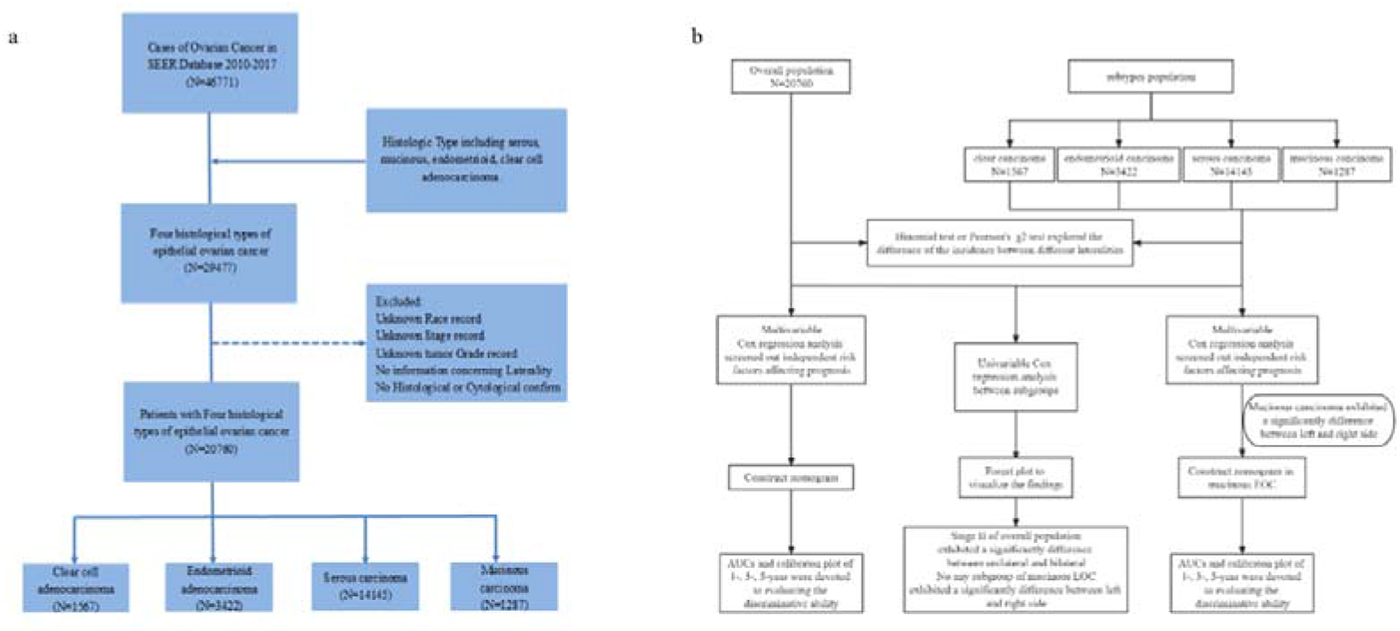
a. Flowchart of datasets with EOC and subtypes, as well as the inclusion criteria and the exclusion criteria. b. Flowchart of statistical analysis in this study. EOC, epithelial ovarian cancer. AUC, the area under the receiver operating characteristic curves.

### 3. Statistical analysis

Binomial test and Pearson’s χ2 test were performed to explore the difference of the incidence between different lateralities. Univariate Cox regression analysis was conducted to examine the impact of independent variables on the prognosis of EOC. Multivariate Cox regression analysis was employed to construct a proportional hazards model for tumour prognosis, and independent risk factors were evaluated with a significance level set at *p* < 0.05. Survival curves were generated using the K-M method to compare the differences in survival between different groups. Depending on the results of the proportional hazards assumption, either Cox analysis or the log-rank test was applied to assess the significance of the survival differences between groups.

### 4. Construction and evaluation of prognostic risk model

Based on the independent risk factors for overall survival (OS) identified through multivariate Cox regression analysis, nomograms were constructed using R software (version 4.2.1). The concordance index (C-index) was utilized to evaluate the accuracy of the nomogram models. A higher C-index indicates a more accurate predictive model. The area under the receiver operating characteristic curve (AUC) was used to assess the discriminative ability of the model. A higher AUC value (closer to 1) indicates better predictive performance of the model. The bootstrap method was employed for internal verification through 200 repeated samplings, and a calibration plot was produced. A higher level of agreement between the predicted and actual survival rates indicates a higher level of model compliance.

## Results

### 1. Lateral incidence comparisons

#### 1. The overall distribution pattern of laterality for EOC

20,760 patients with EOC were included in the study in total, including 5965 (28.7%) patients with left EOC, 6097 (27.4%) patients with right EOC, and 8698 (41.9%) patients with both ovaries. The incidence of unilateral EOC is significantly higher than the one of bilateral in the overall population (unilateral vs. bilateral = 12062(58.1%) vs. 8698(41.9%), *p* < 0.001). Among the unilateral EOC, 5965 patients (49.5%) were left and 6097 patients (50.5%) were right, indicating that there is no significant difference in the overall incidence rate between the left and right sides. However, there were significant differences between the left and right incidence at different stages (*p* = 0.002). Further pairwise comparison indicates that this difference was mainly observed in stage III (Left 47.2% vs Right 52.8%) and stage II (Left 52.2% vs Right 47.8%). (Table 1)

We also characterized the distribution patterns of four subtypes of EOC in the overall population. The results revealed that unilateral tumor pathogenesis predominated in patients with clear cell (unilateral vs bilateral = 1385 (88.4%) vs 182 (11.6%), p < 0.001), endometrioid (unilateral vs bilateral = 2892 (84.5%) vs 530 (15.5%), p < 0.001), and mucinous ovarian cancers (unilateral vs bilateral = 1483 (91.2%) vs 143 (8.8%), *p* < 0.001), whereas bilateral involvement was more common in serous ovarian cancer (unilateral vs bilateral = 6302 (44.6%) vs 7843 (55.4%), *p* < 0.001) (Table 1).

**Table 1.**
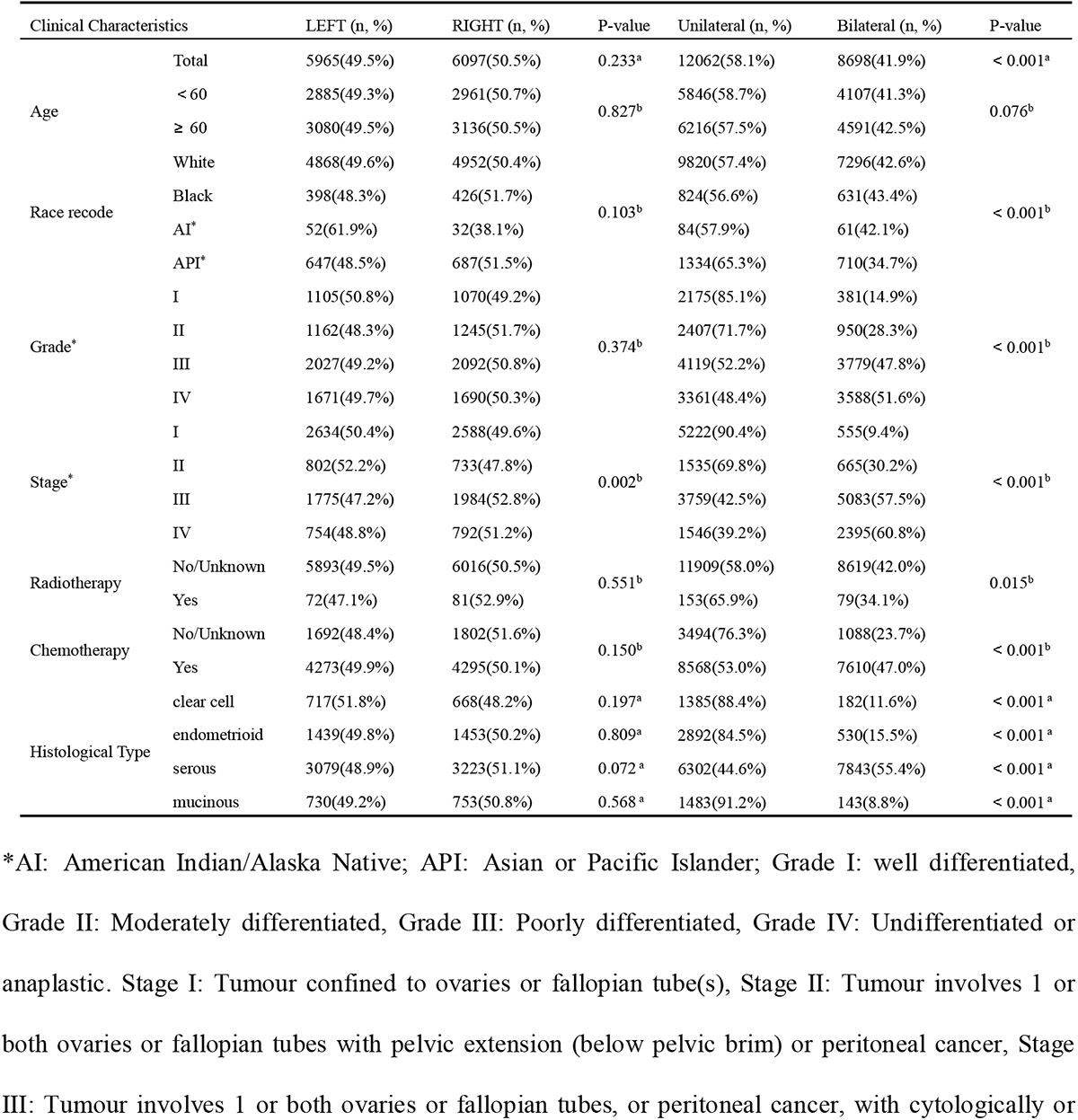
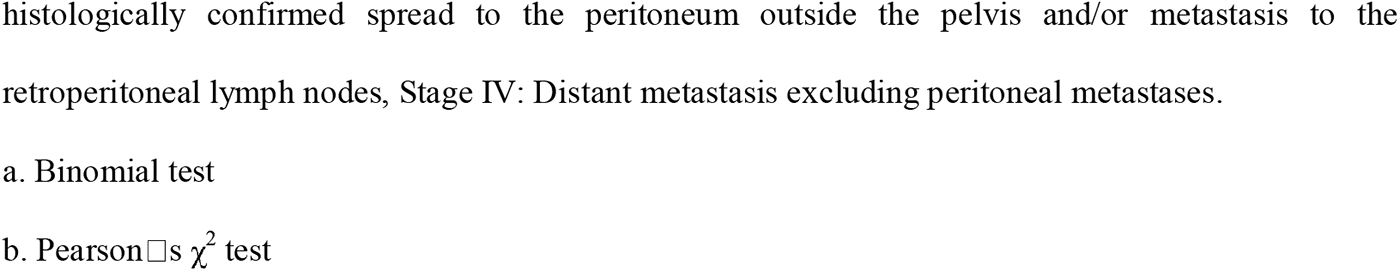
Clinical characteristics and the overall distribution pattern of laterality for epithelial ovarian cancer (EOC)

| Clinical Characteristics |  | LEFT (n, %) | RIGHT (n, %) | P-value | Unilateral (n, %) | Bilateral (n, %) | P-value |
| --- | --- | --- | --- | --- | --- | --- | --- |
| Age | Total | 5965(49.5%) | 6097(50.5%) | 0.233<sup>a</sup> | 12062(58.1%) | 8698(41.9%) | < 0.001<sup>a</sup> |
|  | < 60 | 2885(49.3%) | 2961(50.7%) | 0.827<sup>b</sup> | 5846(58.7%) | 4107(41.3%) | 0.076<sup>b</sup> |
|  | ≥ 60 | 3080(49.5%) | 3136(50.5%) |  | 6216(57.5%) | 4591(42.5%) |  |
| Race recode | White | 4868(49.6%) | 4952(50.4%) | 0.103<sup>b</sup> | 9820(57.4%) | 7296(42.6%) | < 0.001<sup>b</sup> |
|  | Black | 398(48.3%) | 426(51.7%) |  | 824(56.6%) | 631(43.4%) |  |
|  | AI* | 52(61.9%) | 32(38.1%) |  | 84(57.9%) | 61(42.1%) |  |
|  | API† | 647(48.5%) | 687(51.5%) |  | 1334(65.3%) | 710(34.7%) |  |
| Grade* | I | 1105(50.8%) | 1070(49.2%) | 0.374<sup>b</sup> | 2175(85.1%) | 381(14.9%) | < 0.001<sup>b</sup> |
|  | II | 1162(48.3%) | 1245(51.7%) |  | 2407(71.7%) | 950(28.3%) |  |
|  | III | 2027(49.2%) | 2092(50.8%) |  | 4119(52.2%) | 3779(47.8%) |  |
|  | IV | 1671(49.7%) | 1690(50.3%) |  | 3361(48.4%) | 3588(51.6%) |  |
| Stage* | I | 2634(50.4%) | 2588(49.6%) | 0.002<sup>b</sup> | 5222(90.4%) | 555(9.4%) | < 0.001<sup>b</sup> |
|  | II | 802(52.2%) | 733(47.8%) |  | 1535(69.8%) | 665(30.2%) |  |
|  | III | 1775(47.2%) | 1984(52.8%) |  | 3759(42.5%) | 5083(57.5%) |  |
|  | IV | 754(48.8%) | 792(51.2%) |  | 1546(39.2%) | 2395(60.8%) |  |
| Radiotherapy | No/Unknown | 5893(49.5%) | 6016(50.5%) | 0.551<sup>b</sup> | 11909(58.0%) | 8619(42.0%) | 0.015<sup>b</sup> |
|  | Yes | 72(47.1%) | 81(52.9%) |  | 153(65.9%) | 79(34.1%) |  |
| Chemotherapy | No/Unknown | 1692(48.4%) | 1802(51.6%) | 0.150<sup>b</sup> | 3494(76.3%) | 1088(23.7%) | < 0.001<sup>b</sup> |
|  | Yes | 4273(49.9%) | 4295(50.1%) |  | 8568(53.0%) | 7610(47.0%) |  |
| Histological Type | clear cell | 717(51.8%) | 668(48.2%) | 0.197<sup>a</sup> | 1385(88.4%) | 182(11.6%) | < 0.001<sup>a</sup> |
|  | endometrioid | 1439(49.8%) | 1453(50.2%) | 0.809<sup>a</sup> | 2892(84.5%) | 530(15.5%) | < 0.001<sup>a</sup> |
|  | serous | 3079(48.9%) | 3223(51.1%) | 0.072<sup>a</sup> | 6302(44.6%) | 7843(55.4%) | < 0.001<sup>a</sup> |
|  | mucinous | 730(49.2%) | 753(50.8%) | 0.568<sup>a</sup> | 1483(91.2%) | 143(8.8%) | < 0.001<sup>a</sup> |

\*AI: American Indian/Alaska Native; API: Asian or Pacific Islander; Grade I: well differentiated, Grade II: Moderately differentiated, Grade III: Poorly differentiated, Grade IV: Undifferentiated or anaplastic. Stage I: Tumour confined to ovaries or fallopian tube(s), Stage II: Tumour involves 1 or both ovaries or fallopian tubes with pelvic extension (below pelvic brim) or peritoneal cancer, Stage III: Tumour involves 1 or both ovaries or fallopian tubes, or peritoneal cancer, with cytologically or
histologically confirmed spread to the peritoneum outside the pelvis and/or metastasis to the retroperitoneal lymph nodes, Stage IV: Distant metastasis excluding peritoneal metastases.
a. Binomial test
b. Pearson's $\chi^2$ test

#### 2. The distribution pattern of laterality for different subtypes of EOC

The distribution patterns of laterality in the four subtypes of EOC were further characterized, considering the varied clinical characteristics and molecular mechanisms of each subtype (Table 2). In clear cell ovarian cancer, the difference in incidence between unilateral and bilateral cases was primarily observed among different stages, while the distinction between left and right incidence was mainly observed among different races. For both endometrioid and serous ovarian cancers, the primary disparity lied in the incidence rates between unilateral and bilateral cases, as significant differences were observed in almost all subgroups, except for patients who underwent radiotherapy. On the other hand, the discrepancy between left and right incidence were primarily evident among different stages. In mucinous ovarian cancer, there were no significant differences in the incidence rates between the left and right sides among different populations.

**Table 2.** Lateral incidence comparisons in four subtypes of epithelial ovarian cancer (EOC)

|  | LEFT (n, %) | RIGHT (n, %) | P-value | Unilateral (n, %) | Bilateral (n, %) | P-value |
| --- | --- | --- | --- | --- | --- | --- |
| Total |  |  |  |  |  |  |
| Clear cell |  |  |  |  |  |  |
| Age < 60 | 435(53.1%) | 384(46.9%) | 0.228<sup>a</sup> | 819(87.3%) | 119(12.7%) | 0.106<sup>a</sup> |
| Age ≥ 60 | 282(49.8%) | 284(50.2%) |  | 566(90.0%) | 63(10.0%) |  |
| White | 549(53.3%) | 481(46.7%) | 0.034<sup>b</sup> | 1030(88.9%) | 128(11.1%) | 0.321<sup>b</sup> |
| Black | 23(37.7%) | 38(62.3%) |  | 61(82.4%) | 13(17.6%) |  |
| AI* | 6(75.0%) | 2(25.0%) |  | 8(88.9%) | 1(11.1%) |  |
| API* | 139(48.6%) | 147(51.4%) |  | 286(87.7%) | 40(12.3%) |  |
| Grade I | 8(44.4%) | 10 (55.6%) | 0.915<sup>a</sup> | 18(94.7%) | 1(5.3%) | 0.154<sup>b</sup> |
| Grade II | 74(51.7%) | 69(48.3%) |  | 143(92.9%) | 11(7.1%) |  |
| Grade III | 375(52.3%) | 342(47.7%) |  | 717(87.0%) | 107(13.0%) |  |
| Grade IV | 260(51.3%) | 247(48.7%) |  | 507(88.9%) | 63(11.1%) |  |
| Stage I | 477(52.7%) | 428(47.3%) |  | 905(96.6%) | 32(3.4%) |  |
| Stage II | 89(52.7%) | 80(47.3%) | 0.564<sup>a</sup> | 169(88.5%) | 22(11.5%) | □0.001<sup>a</sup> |
| Stage III | 115(49.6%) | 117(50.4%) |  | 232(72.0%) | 90(28.0%) |  |
| Stage IV | 36(45.6%) | 43(54.4%) |  | 79(67.5%) | 38(32.5%) |  |
| No/Unknown Radiotherapy | 703(51.7%) | 657(48.3%) | 0.669<sup>a</sup> | 1360(88.4%) | 179(11.6%) | 1.000<sup>c</sup> |
| Radiotherapy treated | 14(56.0%) | 11(44.0%) |  | 25(89.3%) | 3(10.7%) |  |
| No/Unknown Chemotherapy | 143(51.1%) | 137(48.9%) | 0.794<sup>a</sup> | 280(91.2%) | 27(8.8%) | 0.085<sup>a</sup> |
| Chemotherapy treated | 574(51.9%) | 531(48.1%) |  | 1105(87.7%) | 115(12.3%) |  |
| Endometrioid |  |  |  |  |  |  |
| Age<60 | 879(49.5%) | 898(50.5%) | 0.691<sup>a</sup> | 1777(82.7%) | 372(17.3%) | <0.001<sup>a</sup> |
| Age≥60 | 560(50.2%) | 555(49.8%) |  | 1115(87.6%) | 158(12.4%) |  |
| White | 1190(50.1%) | 1183(49.9%) |  | 2373(85.2%) | 412(14.8%) |  |
| Black | 65(45.5%) | 78(54.5%) | 0.102<sup>a</sup> | 143(76.9%) | 43(23.1%) | 0.019<sup>b</sup> |
| AI* | 14(73.7%) | 5(26.3%) |  | 19(79.2%) | 5(20.8%) |  |
| API* | 170(47.6%) | 187(52.4%) |  | 357(83.6%) | 70(16.4%) |  |
| Grade I | 609(51.0%) | 584(49.0%) |  | 1193(91.5%) | 111(8.5%) |  |
| Grade II | 520(48.6%) | 550(51.4%) | 0.620<sup>a</sup> | 1070(84.9%) | 190(15.1%) | □0.001<sup>a</sup> |
| Grade III | 258(48.8%) | 271(51.2%) |  | 529(72.6%) | 200(27.4%) |  |
| Grade IV | 52(52.0%) | 48(48.0%) |  | 100(77.5%) | 29(22.5%) |  |
| Stage I | 989(49.1%) | 1024(50.9%) |  | 2013(92.0%) | 176(8.0%) |  |
| Stage II | 247(53.8%) | 212(46.2%) | 0.043<sup>a</sup> | 459(79.5%) | 118(20.5%) | □0.001<sup>a</sup> |
| Stage III | 148(45.5%) | 177(54.5%) |  | 325(66.2%) | 166(33.8%) |  |
| Stage IV | 55(57.9%) | 40(42.1%) |  | 95(57.6%) | 70(42.4%) |  |
| No/Unknown Radiotherapy | 1411(49.8%) | 1423(50.2%) | 0.820<sup>a</sup> | 2834(84.7%) | 513(15.3%) | 0.082<sup>a</sup> |
| Radiotherapy treated | 28(48.3%) | 30(51.7%) |  | 58(77.3%) | 17(22.7%) |  |
| No/Unknown Chemotherapy | 567(47.7%) | 622(52.3%) | 0.063<sup>a</sup> | 1189(90.6%) | 124(9.4%) | □0.001<sup>a</sup> |
| Chemotherapy treated | 872(51.2%) | 831(48.8%) |  | 1703(80.7%) | 406(19.3%) |  |
| Serous |  |  |  |  |  |  |
| Age<60 | 1073(48.3%) | 1147(51.7%) | 0.539<sup>a</sup> | 2220(38.6%) | 3529(61.4%) | <0.001<sup>a</sup> |
| Age≥60 | 2006(49.1%) | 2076(50.9%) |  | 4082(48.6%) | 4314(51.4%) |  |
| White | 2552(48.8%) | 2680(51.2%) |  | 5232(44.0%) | 6646(56.0%) |  |
| Black | 254(50.2%) | 252(49.8%) | 0.774<sup>a</sup> | 506(47.6%) | 557(52.4%) | 0.050<sup>a</sup> |
| AI* | 27(54.0%) | 23(46.0%) |  | 50(47.6%) | 55(52.4%) |  |
| API* | 246(47.9%) | 268(52.1%) |  | 514(46.8%) | 585(53.2%) |  |
| Grade I | 149(51.0%) | 143(49.0%) |  | 292(56.2%) | 228(43.8%) |  |
| Grade II | 285(48.6%) | 302(51.4%) | 0.787<sup>a</sup> | 587(45.8%) | 696(54.2%) | □0.001<sup>a</sup> |
| Grade III | 1315(48.3%) | 1407(51.7%) |  | 2722(44.3%) | 3428(55.7%) |  |
| Grade IV | 1330(49.2%) | 1371(50.8%) |  | 2701(43.6%) | 3491(56.4%) |  |
| Stage I | 559(51.6%) | 525(48.4%) |  | 1084(77.3%) | 319(22.7%) |  |
| Stage II | 430(51.4%) | 406(48.6%) | 0.041<sup>a</sup> | 836(62.6%) | 499(37.4%) | □0.001<sup>a</sup> |
| Stage III | 1455(47.3%) | 1618(52.7%) |  | 3073(39.2%) | 4771(60.8%) |  |
| Stage IV | 635(48.5%) | 674(51.5%) |  | 1309(36.7%) | 2254(63.3%) |  |
| No/Unknown Radiotherapy | 3052(48.9%) | 3193(51.1%) | 0.821<sup>a</sup> | 6245(44.5%) | 7785(55.5%) | 0.278<sup>a</sup> |
| Radiotherapy treated | 27(47.4%) | 30(52.6%) |  | 57(49.6%) | 58(50.4%) |  |
| No/Unknown Chemotherapy | 537(47.5%) | 594(52.5%) | 0.306<sup>a</sup> | 1131(55.8%) | 897(44.2%) | □0.001<sup>a</sup> |
| Chemotherapy treated | 2542(49.2%) | 2629(50.8%) |  | 5171(42.7%) | 6946(57.3%) |  |
| Mucinous |  |  |  |  |  |  |
| Age<60 | 498(48.3%) | 532(51.7%) | 0.309<sup>a</sup> | 1030(28.2%) | 87(19.8%) | 0.034<sup>a</sup> |
| Age≥60 | 232(51.2%) | 221(48.8%) |  | 453(30.0%) | 56 (22.1%) |  |
| White | 577(48.7%) | 608(51.3%) |  | 1185(91.5%) | 110(8.5%) |  |
| Black | 56(49.1%) | 58(50.9%) | 0.573<sup>b</sup> | 114(86.4%) | 18(13.6%) | 0.179<sup>a</sup> |
| AI<sup>*</sup> | 5(71.4%) | 2(28.6%) |  | 7(100.0%) | 0(0.0%) |  |
| API<sup>*</sup> | 92(52.0%) | 85(48.0%) |  | 177(92.2%) | 15(7.8%) |  |
| Grade I | 339(50.4%) | 333(49.6%) |  | 672(94.2%) | 41(5.8%) |  |
| Grade II | 283(46.6%) | 324(53.4%) | 0.353<sup>a</sup> | 607(92.0%) | 53(8.0%) | □0.001<sup>a</sup> |
| Grade III | 79(52.3%) | 72(47.7%) |  | 151(77.4%) | 44(22.6%) |  |
| Grade IV | 29(54.7%) | 24(45.3%) |  | 53(91.4%) | 5(8.6%) |  |
| Stage I | 609(49.9%) | 611(50.1%) |  | 1220(92.8%) | 28(2.2%) |  |
| Stage II | 36(50.7%) | 35(49.3%) | 0.535<sup>a</sup> | 71(73.2%) | 26(26.8%) | □0.001<sup>a</sup> |
| Stage III | 57(44.2%) | 72(55.8%) |  | 129(69.7%) | 56(30.3%) |  |
| Stage IV | 28(44.4%) | 35(44.6%) |  | 63(65.6%) | 33(34.4%) |  |
| No/Unknown Radiotherapy | 727(49.5%) | 743(50.5%) | 0.058<sup>a</sup> | 1470(91.2%) | 142(8.8%) | 1.000<sup>c</sup> |
| Radiotherapy treated | 3(23.1%) | 10(76.9%) |  | 13(92.9%) | 1(7.1%) |  |
| No/Unknown Chemotherapy | 445(49.8%) | 449(50.2%) | 0.601<sup>a</sup> | 894(95.7%) | 40(4.3%) | □0.001<sup>a</sup> |
| Chemotherapy treated | 285(48.4%) | 304(51.2%) |  | 589(91.2%) | 103 (8.8%) |  |

### 2. Prognosis evaluation

#### 2.1 Bilateral had a worse prognosis than unilateral EOC in the overall population

Age, ethnicity, laterality (left, right and bilateral), FIGO stage, tumour grade and whether there was chemotherapy and radiotherapy were included as independent prognostic factors. Multivariate cox regression analysis showed that older age (HR = 1.526, *p* < 0.001), bilateral tumours (HR = 1.114, *p* < 0.001), black race (HR = 1.271, *p* < 0.001), higher grade and higher stage and radiotherapy (HR = 1.435, *p* < 0.001) were independent risk factors for EOC, and chemotherapy (HR = 0.534, *p* < 0.001) was independent favourable factors for EOC (Table 3). For the variable of laterality, there was no significant difference in prognosis between patients with right-sided and left-sided EOC, with a hazard ratio of 0.987 (*p* = 0.692, 95%CI: 0.925-1.053) (Table 2). Patients with bilateral ovarian cancer, however, had a worse prognosis than those with unilateral ovarian cancer, with a hazard ratio of 1.120 (*p* < 0.001, 95%CI: 1.070-1.173).

**Table 3.**
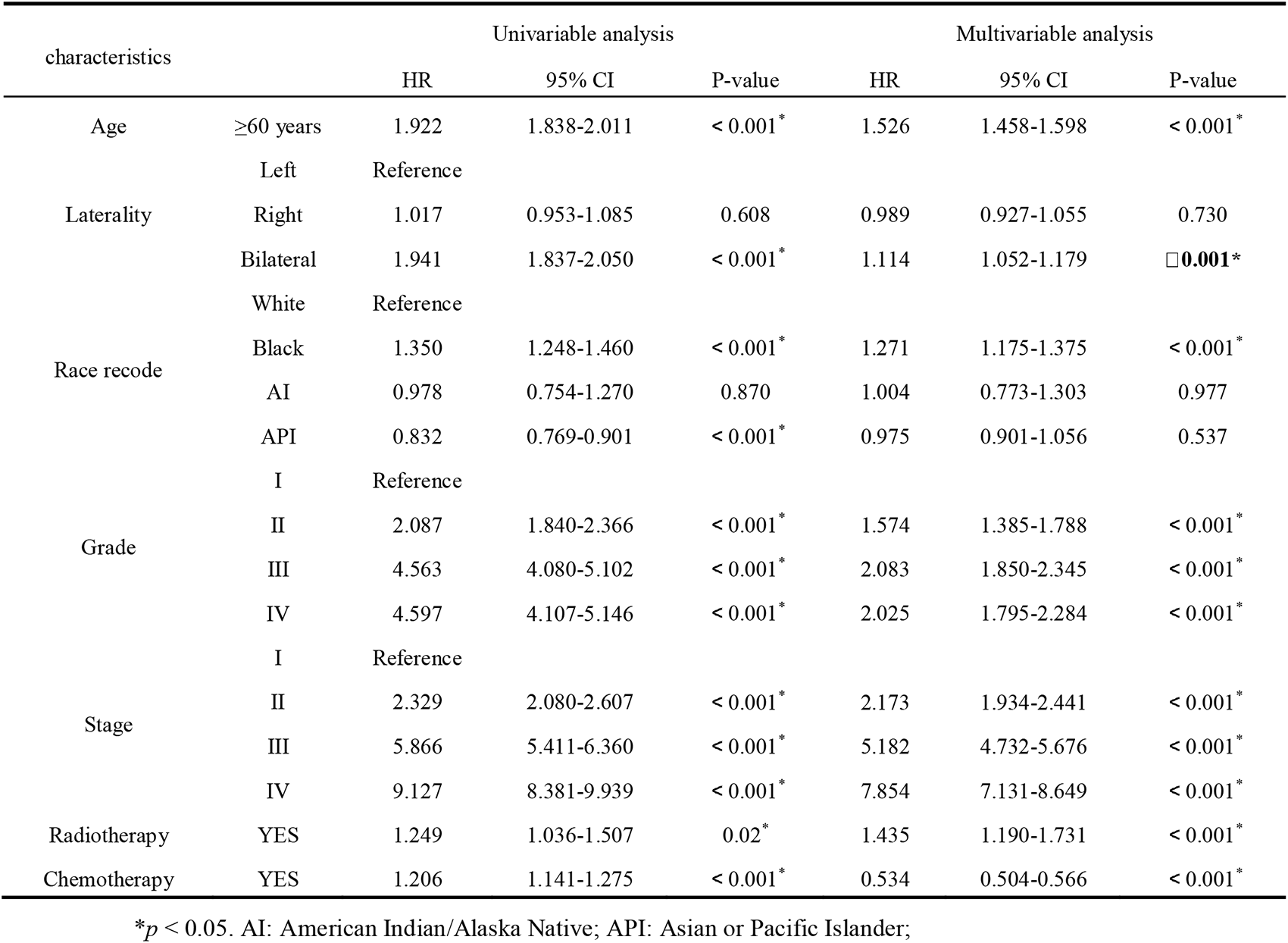
Univariable and multivariable analyses of clinicopathological characteristics as predictors of overall survival (OS) for epithelial ovarian cancer (EOC).

We further divided EOC patients into different subgroups according to laterality, i.e., unilateral vs. bilateral. The KM plot was used to compare survival differences between groups. Again, results showed that the unilateral EOC had a significant prognostic advantage over bilateral EOC (*p* < 0.0001) (Figure 2).

**Figure 2.**
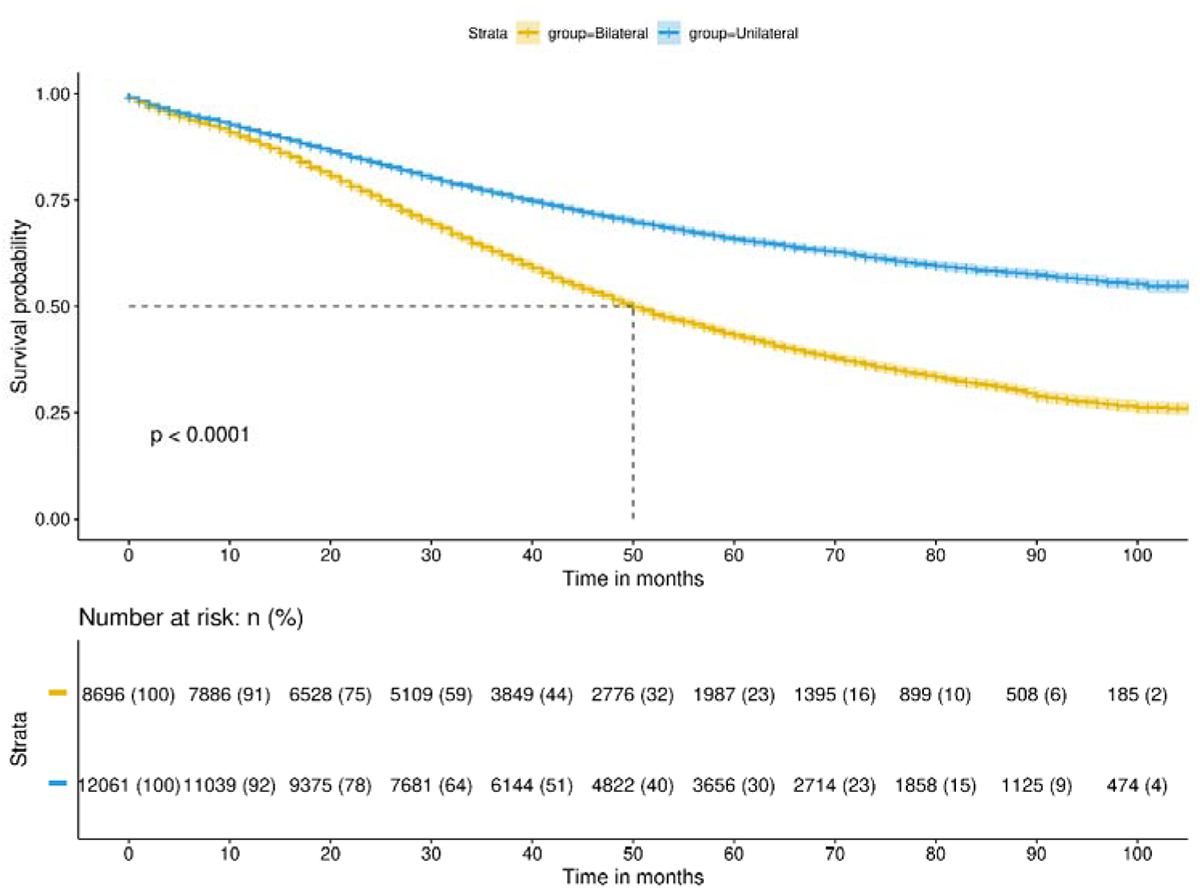
Unilateral vs. bilateral Kaplan-Meier survival curves in overall population. The median survival for bilateral tumours is 50 months, while the median survival for unilateral tumours is over 100 months and still not reached.

Independent risk factors filtrated based on multivariate cox regression analysis were used to construct nomograms for 1-, 3- and 5-year OS (Figure 3a). The C-index of the nomogram was 0.724 [95% confidence interval (CI): 0.718−0.730]. The AUCs of the 1-, 3-, and 5-year nomogram models were 0.751, 0.752, and 0.775, respectively. The calibration plots all demonstrated good agreement between the predictive values of OS for 1-, 3- and 5-year survival and the actual survival probability (Figure 3b, Figure 3c).

**Figure 3.**
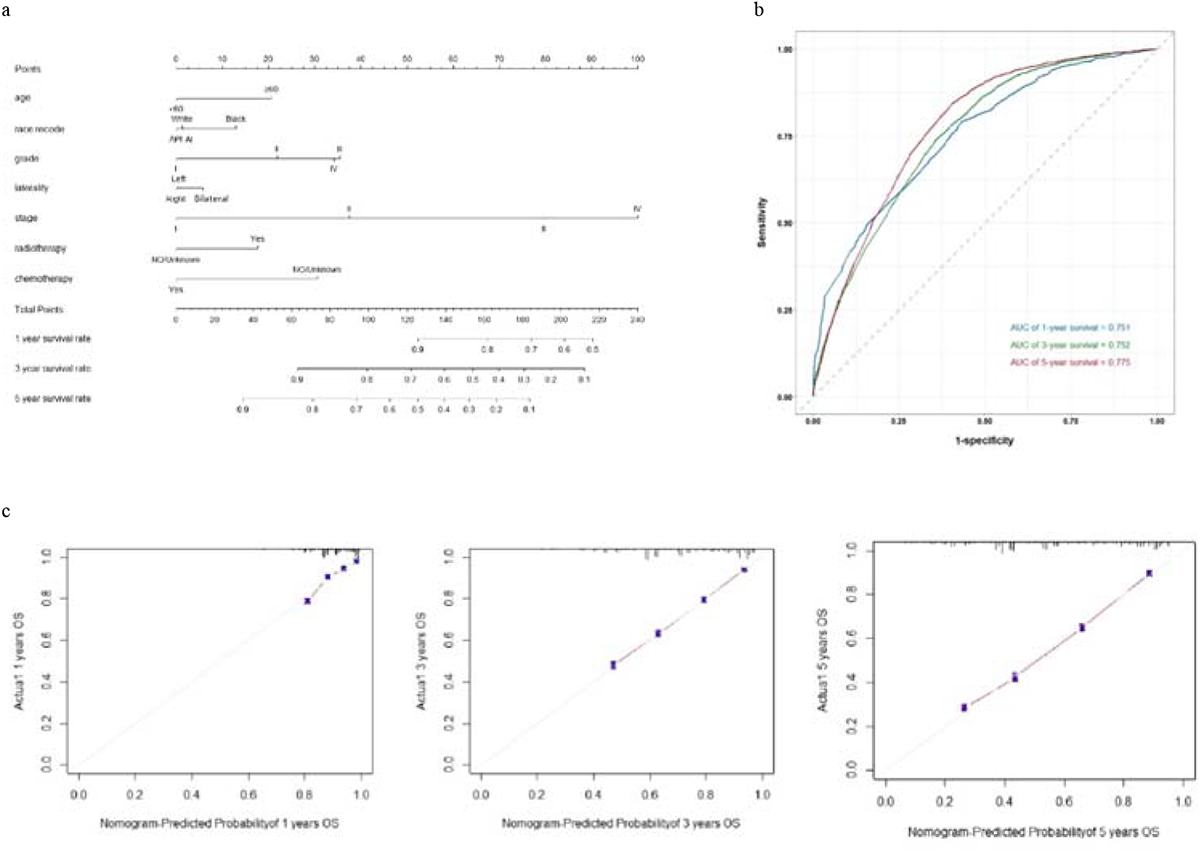
a. Based on the independent risk factors screened by multivariate cox regression analysis of the overall population, a nomogram was drawn to predict the survival rate of 1-, 3-, and 5-years respectively. b. The area under the receiver operating characteristic curves (AUCs) of the 1-, 3-, and 5-year were devoted to evaluating the discriminative ability of the nomogram models. c. The calibration plots of OS for 1-, 3- and 5-year.

#### 2.2 Bilateral EOC has a significantly worse prognosis than unilateral EOC, especially in stage II

When we focused on the prognosis effects of laterality on subgroup population divided by significant variables obtained from multivariate cox regression analysis above, bilateral EOC exhibited a significantly worse prognosis than unilateral EOC in most subgroups, except advanced EOC (Stage III/IV). Especially in stage II, that bilateral EOC has a significantly worse prognosis than unilateral EOC which is not mentioned in the FIGO staging system^27^. To account for multiple testing, two-sided *p* values were adjusted according to the method of Benjamini/Hochberg (B/H) to control the false discovery rate (FDR). An association was considered to be statistically significant, if its corresponding B/H-adjusted p value was below 0.05, corresponding to an FDR of 5%. (Figure 4).

**Figure 4.**
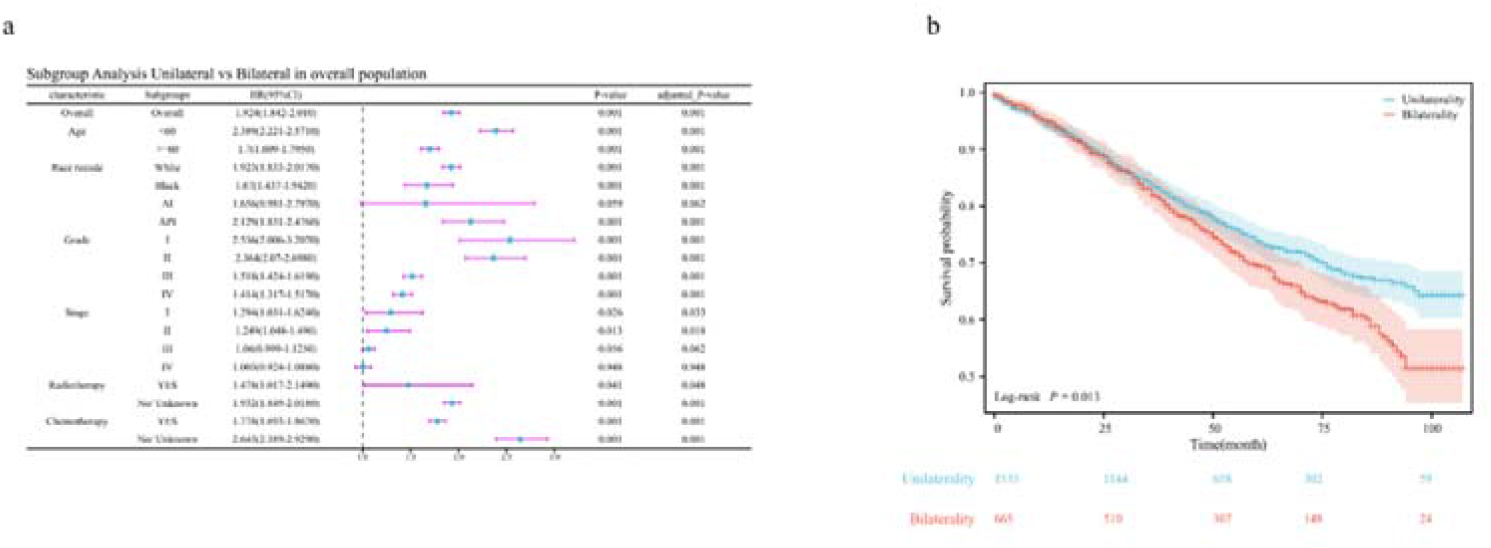
a. Subgroup analysis Unilateral vs. Bilateral in overall population; The last two columns list the unadjusted and adjusted p values according to Benjamini/Hochberg (BH), respectively. All 0.001 refers to less than 0.001.b. Survival curve between Unilateral and Bilateral in Stage II.

#### 2.3 The prognosis of right-side is worse than the left-side in mucinous ovarian cancer

In each subtype of histology, the effect of laterality on tumour prognosis was mostly consistent with the effect in the overall population we identified above (Table 3), that is patients with left and right ovarian cancer were nearly overlap with no statistical significance, but the unilateral EOC has a significant prognostic advantage over bilateral EOC (Table 4 and Supplementary File 1). However, in patients with mucinous ovarian cancer, the right-to-left prognostic hazard ratio was 0.745 (*p* = 0.015, 95%CI 0.587 to 0.945) (Table 4), indicating that right mucinous ovarian cancer exhibited a better prognosis than left.

**Table 4.**
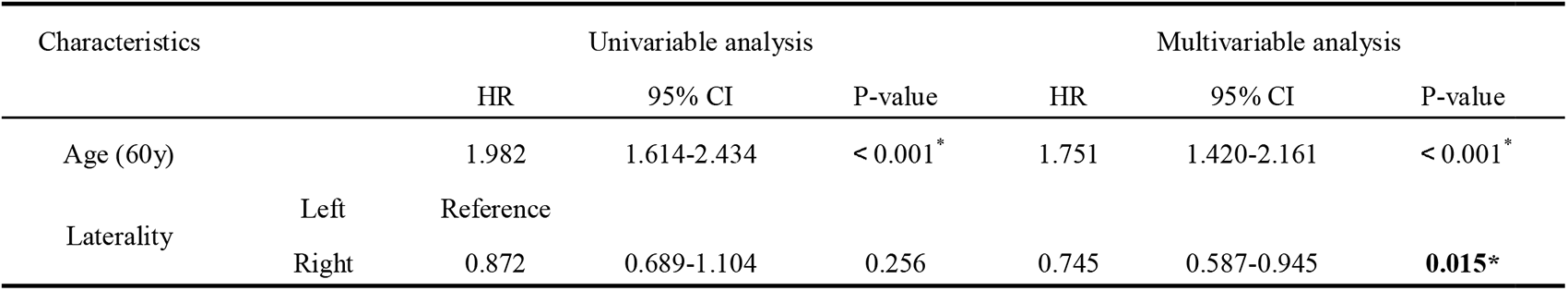
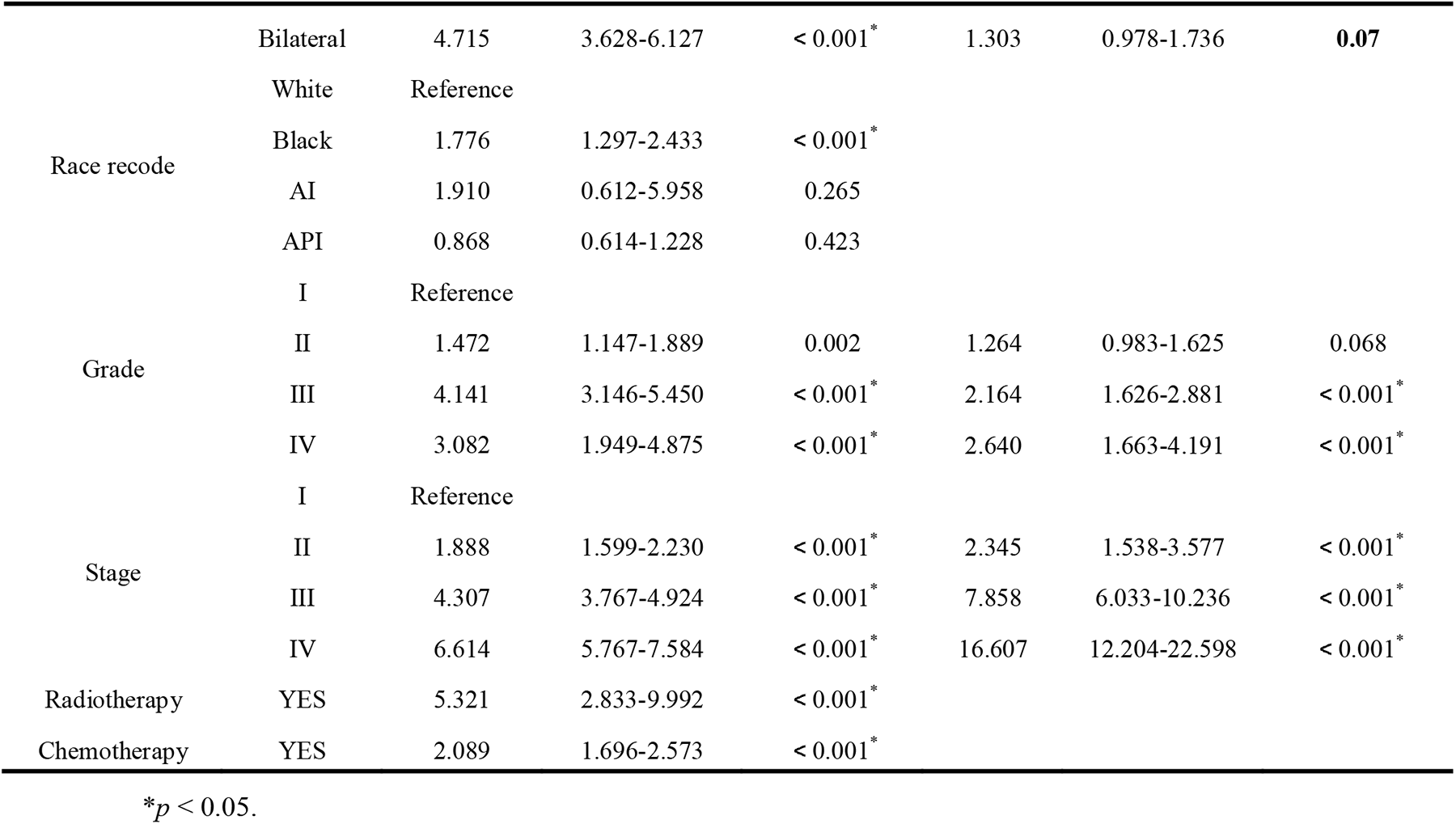
Univariable and multivariable analyses of overall survival (OS) for mucinous ovarian cancer.

Based on the result of COX proportional hazards model of mucinous ovarian cancer, we construct the nomogram (Fig 5a). The C-index of the predictive OS nomogram of mucinous ovarian cancer was 0.818 [95% confidence interval (CI): 0.794−0.842]. The AUCs of the 1-, 3-, and 5-year nomogram models were 0.870, 0.850, and 0.815, respectively. The calibration plots also demonstrated excellent agreement between the predictive values of OS for 1-, 3- and 5-year survival and the actual survival probability (Fig 5b, Fig 5c). This predictive OS nomogram of mucinous ovarian cancer shows higher consistency than of overall population. And the AUCs were closer to 1, showing good prediction performance.

**Figure 5.**
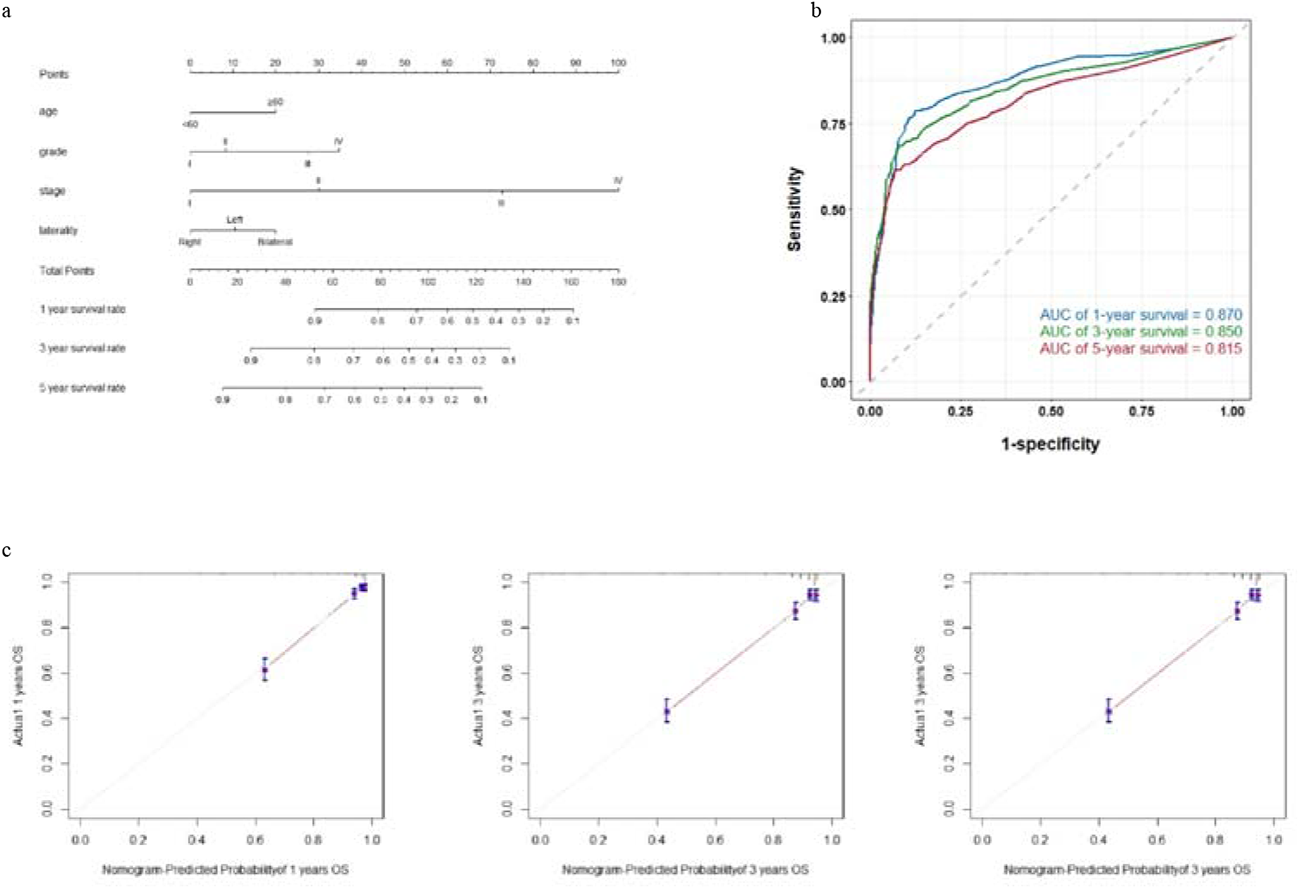
a. Based on the independent risk factors screened by multivariate cox regression analysis of the mucinous ovarian cancer, a nomogram was drawn to predict the survival rate of 1-, 3-, and 5-years respectively. b. The area under the receiver operating characteristic curves (AUCs) of the 1-, 3-, and 5-year were devoted to evaluating the discriminative ability of the nomogram models. c. The calibration plots of OS for 1-, 3- and 5-year.

## Discussion

This study investigated the impact of ovarian laterality (unilateral vs. bilateral, left-side vs. right-side) on both incidence and prognosis of EOC in the overall population and among different subtypes. In the overall population, the laterality difference primarily occurred between unilateral and bilateral cases. The bilateral incidence rate was lower than that of unilateral cases, and bilateral prognosis was worse than unilateral prognosis in the overall population, which are as expected. Notably, the impact of laterality on incidence rate varied among different EOC subtypes. Serous ovarian cancer predominantly occurred bilaterally, while the other three subtypes primarily occurred on the unilateral side, consistent with previous reports. Initially, we hypothesized that this difference may be due to the bilateral origins of serous ovarian cancer. However, genetic studies on bilateral invasive ovarian tumours have shown that the majority of cases of serous ovarian cancer are monoclonal, representing spread from a single ovarian site^28^. Then we proposed that the discrepancy could be attributed to the varying frequency of different subtypes at the initial diagnosis stage. Our dataset revealed that most serous ovarian cancer cases were diagnosed at advanced stages, such as stage III (51%) or IV (29%). This aligns with clinical practice, as serous ovarian cancer is typically challenging to detect at an early stage, due to its small size at the early stages, making it difficult to develop symptoms without metastasis or complications. Additionally, serous ovarian cancer cells are mostly high-grade, type II epithelial, which are aggressive and cause the disease to progress rapidly^5^. Consequently, serous ovarian cancer are often detected at advanced stages, which could explain why it is predominantly detected bilaterally in our study. In contrast, the majority (58% to 64%) of endometrioid, mucinous, and clear cell ovarian cancer cases in the dataset were diagnosed at earlier stages, such as stage I^16,29^. This can be attributed to the fact that endometrioid and clear cell ovarian cancers are endometriosis-related diseases, which are relatively easier to detect early due to symptoms such as dysmenorrhea. Additionally, the larger size of mucinous ovarian cancer also facilitates early diagnosis compared to serous ovarian cancer^30^.

It is noteworthy that the incidence rate between the left and right sides significantly varied among different stages of EOC in the overall population. Furthermore, significant differences were also observed in the incidence rates between the left and right sides, as well as between unilateral and bilateral cases among different stages in many EOC subtypes. Moreover, we identified stage as an important factor influencing the disparities in prognosis between unilateral and bilateral cases. All these results consistently demonstrated a significant relationship between stage and the differences in lateral incidence rates. As we know that tumours at different stages often have distinct genetic characteristics and clinical features^5^. The FIGO staging system is widely used for gynaecological malignancies, including EOC, but it does not adequately consider the laterality of the ovary, except in stage I where unilateral or bilateral involvement is considered as a prognostic factor^27^. Our study revealed that ovarian laterality, both for unilateral-bilateral and left-right comparisons, has a significant impact on both incidence and prognosis of EOC at different stages.

The impact of laterality on the prognosis of different EOC subtypes also varies, particularly for mucinous ovarian cancer. We observed that the prognosis on the right side was significantly better than that on the left side for mucinous ovarian cancer. While univariate Cox regression analysis revealed no significant difference in prognosis between the left and right sides, multivariate Cox regression analysis demonstrated a significant difference (*p* < 0.05). The inconsistency between these results may be attributed to the correlation between other confounding factors and laterality in the univariate analysis, which masks the true effect of laterality. However, multivariate analysis accounted for this effect by including other variables, revealing laterality as an independent factor affecting mucinous ovarian cancer prognosis. To the best of our knowledge, no literature has reported this phenomenon.

The reasons for the poorer prognosis on the left side compared to the right side of mucinous ovarian cancer require further investigation. Here, we can only offer some inferences based on existing research findings. Primary mucinous ovarian cancer is a rare malignancy, accounting for less than 3% of all EOC cases. Nearly 60% of mucinous ovarian cancer cases are metastatic ovarian tumors, with the stomach or colorectum being the most common origin of metastasis^24,30,31^. The lateral preference for metastasis may be a related factor. Anatomical differences between the left and right hemipelvis could provide an explanation. Charles Chapron et al^32^. indicated that the close anatomical relationship between the sigmoid colon and the left adnexa forms a barrier to the pelvic diffusion of menstrual blood reflux. This has been used to explain why deeply infiltrating endometriosis lesions are more commonly observed in the posterior pelvic compartment and predominantly located on the left side. This asymmetric anatomy of the pelvic cavity may also affect the direct spread and implantation of cancer cells, thus potentially explaining why the prognosis on the left side is poor for mucinous ovarian cancer. Furthermore, variations in venous structures may also be a factor that affects the distant metastasis of cancer cells, consequently influencing the prognosis. Hematogenous metastasis is one of the mechanisms of distant metastasis in ovarian cancer tumors. The right ovarian reflux vein joins the inferior vena cava, while the left ovarian blood flows back into the renal vein^33^. Due to the thinness of the renal vein and its susceptibility to reflux obstruction, pelvic varices in the left ovary are more common^34^. Lastly, differences in the invasive and metastatic capacities of left and right mucinous ovarian cancer cells themselves may also contribute to the disparity in prognosis. Further comprehensive research is necessary to elucidate the underlying mechanisms related to the different prognosis between the left and right sides of mucinous ovarian cancer.

We also found significant effects of age and race on the incidence rates and prognostic differences between unilateral and bilateral cases, as well as between the left and right sides, both in the overall population and across different subtypes. Indeed, studies have reported the effects of age and race on disease incidence rates and prognosis. For instance, Lindsey A. Torre et al^29^. demonstrated that API women have the highest incidence rate of clear cell ovarian cancer and the lowest incidence rate of serous ovarian cancer. Therefore, it is important to consider the impacts of age and race on the laterality of incidence rates and prognosis in clinical practice.

To the best of our knowledge, our study is the first to systematically investigate the impact of laterality on the incidence and prognosis of EOC. Our results suggest that whether the tumour occurs unilaterally or bilaterally, the left-side or right-side, significantly influences the incidence and prognosis of EOC, particularly in different subtypes and stages. These findings should be taken into consideration in clinical practice.

## Data Availability

All data produced in the present work are contained in the manuscript

https://seer.cancer.gov/

## Declarations

### Ethics approval and consent to participate

Not applicable.

### Consent for publication

Not applicable.

### Availability of data and materials

Information about the data used in this study were included in this article and its supplementary files.

### Authors’ Contributions

GY and LG conceived the study. YZ conducted the analyses. YZ, YH and JK conducted data curation. SY and GY gave the funding supports. YZ and LG wrote the manuscript and all authors commented on it.

## Acknowledgements

We thank Prof. Canwei Xia and Prof. Xia Shen for the valuable suggestions.

