## Supplementary File 1 for "The Impact of Laterality on the Incidence and Prognosis of Epithelial Ovarian Cancer"

**Supplementary Table 1** Univariable and multivariable analyses of overall survival (OS) for clear cell ovarian cancer

| characteristics |  | Univariable analysis | | | Multivariable analysis | | |
| --- | --- | --- | --- | --- | --- | --- | --- |
|  |  | HR | 95% CI | P-value | HR | 95% CI | P-value |
| Age (60y) |  | 1.097 | 0.913-1.319 | 0.322 |  |  | 0.979 |
| Laterality | Left | Reference |  |  |  |  |  |
|  | Right | 1.060 | 0.859-1.309 | 0.588 | 1.004 | 0.813-1.242 | 0.968 |
|  | Bilateral | 3.807 | 3.013-4.812 | ＜0.001 | 1.829 | 1.432-2.338 | ＜0.001 |
| Race recode | White | Reference |  |  |  |  |  |
|  | Black | 1.839 | 1.278-2.644 | 0.001 | 1.934 | 1.341-2.791 | ＜0.001 |
|  | AI | 0.368 | 0.052-2.617 | 0.318 | 0.482 | 0.068-3.436 | 0.466 |
|  | API | 1.121 | 0.899-1.399 | 0.311 | 1.117 | 0.894-1.395 | 0.330 |
| Grade | I | Reference |  | 0.013 |  |  |  |
|  | II | 0.778 | 0.303-1.996 | 0.601 |  |  |  |
|  | III | 1.370 | 0.566-3.319 | 0.485 |  |  |  |
|  | IV | 1.154 | 0.474-2.810 | 0.753 |  |  |  |
| Stage | I | Reference |  |  |  |  |  |
|  | II | 2.626 | 1.931-3.572 | ＜0.001 | 2.564 | 1.882-3.493 | ＜0.001 |
|  | III | 6.400 | 5.118-8.003 | ＜0.001 | 5.740 | 4.545-7.248 | ＜0.001 |
|  | IV | 10.208 | 7.718-13.502 | ＜0.001 | 8.833 | 6.608-11.809 | ＜0.001 |
| Radiotherapy |  | 1.608 | 0.884-2.926 | 0.120 |  |  |  |
| Chemotherapy |  | 0.881 | 0.704-1.103 | 0.270 | 0.707 | 0.564-0.887 | 0.003 |

**Supplementary Table 2** Univariable and multivariable analyses of overall survival (OS) for endometrioid ovarian cancer

| characteristics |  | Univariable analysis | | | Multivariable analysis | | | |
| --- | --- | --- | --- | --- | --- | --- | --- | --- |
|  |  | HR | 95% CI | P-value | HR | | 95% CI | P-value |
| Age (60y) |  | 2.308 | 1.959-2.719 | ＜0.001 | 1.777 | 1.503-2.101 | | 0.979 |
| Laterality | Left | Reference |  |  |  |  | |  |
|  | Right | 0.955 | 0.788-1.156 | 0.634 |  |  | |  |
|  | Bilateral | 2.166 | 1.765-2.658 | ＜0.001 |  |  | |  |
| Race recode | White | Reference |  |  |  |  | |  |
|  | Black | 1.482 | 1.088-2.018 | 0.013 |  |  | |  |
|  | AI | 0.653 | 0.210-2.031 | 0.461 |  |  | |  |
|  | API | 0.972 | 0.751-1.257 | 0.826 |  |  | |  |
| Grade | I | Reference |  |  |  |  | |  |
|  | II | 1.937 | 1.521-2.466 | ＜0.001 | 1.647 | 1.286-2.109 | | ＜0.001 |
|  | III | 4.848 | 3.846-6.112 | ＜0.001 | 2.307 | 1.778-2.993 | | ＜0.001 |
|  | IV | 5.749 | 4.044-8.172 | ＜0.001 | 2.832 | 1.948-4.116 | | ＜0.001 |
| Stage | I | Reference |  |  |  |  | |  |
|  | II | 2.053 | 1.603-2.630 | ＜0.001 | 1.985 | 1.540-2.575 | | ＜0.001 |
|  | III | 5.896 | 4.832-7.197 | ＜0.001 | 4.877 | 3.881-6.130 | | ＜0.001 |
|  | IV | 11.064 | 8.645-14.159 | ＜0.001 | 8.816 | 6.719-11.567 | | ＜0.001 |
| Radiotherapy |  | 0.799 | 0.427-1.492 | 0.481 |  |  | |  |
| Chemotherapy |  | 1.100 | 0.929-1.302 | 0.269 | 0.578 | 0.482-0.693 | | ＜0.001 |

**Supplementary Table 3** Univariable and multivariable analyses of overall survival (OS) for serous ovarian cancer

| characteristics |  | Univariable analysis | | | Multivariable analysis | | |
| --- | --- | --- | --- | --- | --- | --- | --- |
|  |  | HR | 95% CI | P-value | HR | 95% CI | P-value |
| Age (60y) |  | 1.674 | 1.592-1.761 | ＜0.001 | 1.580 | 1.501-1.662 | ＜0.001 |
| Laterality | Left | Reference |  |  |  |  |  |
|  | Right | 1.046 | 0.969-1.130 | 0.245 | 1.014 | 0.939-1.095 | 0.723 |
|  | Bilateral | 1.336 | 1.254-1.442 | ＜0.001 | 1.126 | 1.056-1.200 | ＜0.001 |
| Race recode | White | Reference |  |  |  |  |  |
|  | Black | 1.250 | 1.147-1.363 | ＜0.001 | 1.240 | 1.137-1.353 | ＜0.001 |
|  | AI | 1.024 | 0.775-1.352 | 0.870 | 1.058 | 0.801-1.398 | 0.692 |
|  | API | 0.902 | 0.821-0.991 | 0.031 | 0.915 | 0.833-1.005 | 0.065 |
| Grade | I | Reference |  |  |  |  |  |
|  | II | 1.901 | 1.558-2.318 | ＜0.001 | 1.687 | 1.383-2.058 | ＜0.001 |
|  | III | 2.873 | 2.394-3.448 | ＜0.001 | 2.195 | 1.826-2.638 | ＜0.001 |
|  | IV | 2.787 | 2.322-3.345 | ＜0.001 | 2.187 | 1.818-2.631 | ＜0.001 |
| Stage | I | Reference |  |  |  |  |  |
|  | II | 1.888 | 1.599-2.230 | ＜0.001 | 1.883 | 1.593-2.225 | ＜0.001 |
|  | III | 4.307 | 3.767-4.924 | ＜0.001 | 4.416 | 3.849-5.067 | ＜0.001 |
|  | IV | 6.614 | 5.767-7.584 | ＜0.001 | 6.671 | 5.792-7.682 | ＜0.001 |
| Radiotherapy |  | 1.437 | 1.153-1.791 | 0.001 | 1.449 | 1.162-1.807 | ＜0.001 |
| Chemotherapy |  | 0.744 | 0.697-0.793 | ＜0.001 | 0.509 | 0.476-0.544 | ＜0.001 |
